# Flexible and scalable participatory syndromic and virological surveillance for respiratory infections: our experiences in The Netherlands

**DOI:** 10.1101/2024.04.24.24306278

**Authors:** Tara Smit, Gesa Carstens, Wanda Han, Kirsten Bulsink, Jordy de Bakker, Mansoer Elahi, Rianne van Gageldonk-Lafeber, Susan van den Hof, Dirk Eggink, Albert Jan van Hoek

## Abstract

**Background:** During the COVID-19 pandemic participatory digital syndromic surveillance systems proved itself, as it is scalable, flexible and function independent from the health care system or health care seeking behaviour. A limitation of syndromic surveillance is the inability of pathogen identification. We describe our experiences regarding integrating self-swabs with centralized testing into a participatory syndromic surveillance system in the Netherlands (*Infectieradar*).

**Methods:** In the 2022/2023 winter season *Infectieradar* was extended to include nose- and throat swabs. Participants received test-kits including SARS-CoV-2 antigen tests for home use as well as nose- and throat swabs. All SARS-CoV-2 positive participants and a random sample of symptomatic SARS-CoV-2 self-test negative participants were asked to return a nose- and throat swab by regular post. Self-test negative swabs were tested by multiplex-PCR on 22 pathogens, including SARS-CoV-2. Self-test SARS-CoV-2 positive samples with a Ct-value < 30 were sequenced for variant analysis.

**Results:** Over 17,000 participants were included in the study. We collected 1,475 (median: 37 per week) swabs from participants with positive and 4,096 swabs (median: 136 per week) from participants with negative SARS-CoV-2 antigen self-tests. Of the swabs following a negative self-test, 47.7% tested positive in the multiplex-PCR, and rhinovirus/enterovirus was the most frequently detected pathogen (24.5%). Self-test SARS-CoV-2 positivity was laboratory-confirmed in 96.1% of swabs and showed parallel variant distributions as the national SARS-CoV-2 variant surveillance.

**Conclusion:** This large-scale, centralized participatory surveillance system provides a comprehensive approach for performing syndromic and virological surveillance in the general population, including respiratory pathogen detection by self-test or multiplex-PCR. Given the continuous collection of samples among those who don’t seek care, the system provides valuable insights into circulating respiratory pathogens and is part of an answer on how to study the transmission, competition, virulence and evolution of circulating pathogens in interpandemic periods.

## Introduction

Respiratory infectious diseases such as influenza and coronavirus disease 2019 (COVID-19) can be monitored in populations through a variety of surveillance systems. These systems include sentinel surveillance in affiliated general practitioner (GP) practices and hospitals, and non-sentinel systems that gather pathogen-specific testing data from laboratories, which obtain samples from hospitals, GPs and long-term care facilities. Other systems, such as vaccine uptake monitors, sero-epidemiological studies, wastewater analysis, excess mortality monitors, and participatory syndromic surveillance, may also be implemented to broaden and strengthen respiratory infectious disease surveillance systems across the surveillance pyramid. Such systems are crucial for public health decision-making and preparedness for potential emerging diseases (1).

Each surveillance system has its own limitations and strengths depending on their targeted disease severity, population group, and timeliness. Wastewater analysis, for example, provides information on prevalence based on the number of virus particles, including those from both symptomatic and asymptomatic individuals independent of healthcare-seeking behavior. However, this system does not allow for linking to individual-level data such as disease severity, age, sex and clinical risk group. Conversely, GP sentinel surveillance might have information on age, sex and clinical risk group, but relies on health care visits. Visits to GPs can be biased towards individuals at higher risk of adverse health effects, and missing those who are symptomatic but do not seek care. In addition, there are differences in timeliness; the time between infection, viral shedding, symptom onset and a formal diagnosis, or hospitalization and death (2). Therefore, to achieve comprehensive situational awareness, multiple respiratory infectious diseases surveillance systems are required, performing in parallel.

To have a surveillance system independent of care capacity and healthcare seeking behaviour, a digital participatory syndromic surveillance system (*Infectieradar*) was launched in the Netherlands at the beginning of the COVID-19 epidemic (2, 3). *Infectieradar* is a cohort of around 12,000 participants who report their symptoms weekly related to infectious diseases such as COVID-19 and other acute respiratory infections (ARI). This surveillance system informed decision making throughout the pandemic and became increasingly important during the later stages of the pandemic, when SARS-CoV-2 community testing facilities were not available anymore for the general population. Additional advantages of digital surveillance over other surveillance systems (4, 5) include scalability, social, demographic, economic and health background information per case, independence from capacity in testing facilities, easily adaptable questionnaires, and a lower threshold for reporting mild symptoms. However, a limitation of digital participatory syndromic surveillance is the ability of pathogen identification.

To overcome the limitation of absence of pathogen detection in digital surveillance, we integrated self-swabs and central testing for pathogen identification within *Infectieradar* starting in the 2022/2023 respiratory season. To our knowledge, this is the first nationwide participatory syndromic surveillance system in which self-tests are distributed, self-swab samples are collected and analyzed for a broad panel of respiratory pathogens.

Therefore, the objective of this paper is to describe the methods used to include self-tests and self-swabs in our participatory syndromic surveillance system (5) and evaluate its implementation and results regarding both the distribution of SARS-CoV-2 variants and of other pathogens associated with respiratory symptoms.

## Methods

*Infectieradar* was launched in March, 2020. The design and methods have been described in detail (2). Briefly, once registered, participants fill in an intake questionnaire collecting sociodemographic data and medical history data. After that, participants receive a weekly e-mail notification to fill in a weekly questionnaire about whether they experience symptoms of an acute infection. Although each participant is assigned to a specific day and receives the notification that day, they are free to complete the weekly questionnaire at any time and multiple times in the same week.

The testing component was added in the season 2022/2023. We offered participation to our existing users, and additionally recruited new participants. To improve our population representation, we performed random sampling of 300,000 individuals from the Dutch national population registry (known as the Basis Registratie Persoonsgegevens; BRP). To account for an anticipated lower response among young age groups and males, we oversampled these groups, based on the non-response rate observed in a previous recruitment effort in the Netherlands (6). The 300.000 individuals were selected between age 16 (legal minimum age for participating in medical scientific research in the Netherlands) and 74, assuming that individuals within this age range were proficient in using digital devices.

Recruitment invitations were sent between August 31, 2022 and October 3, 2022. Existing users received an invitation email with a hyperlink to a personal login page for enrolment. Individuals identified from the BRP received invitation letters by post, which included a hyperlink and QR code for study enrolment. Each invitation was assigned a unique personal code which was checked upon registration to assure that only those with an invitation could sign-up.

During the registration process, participants were required to provide their first and last names, address, place of residence, zip code, phone number (used for two-factor authentication), and email address. This information is used to run the study, but are stored and curated independently from the research data.

We constrained the maximum number of total participants to 50,000, in line with our logistical capacity to provide self-test packages to participants. Since this number was not reached in our recruitment effort the cohort remained open for self-registrants after the 3^rd^ of October.

Following registration, all enrolled participants received a self-test package. The self-test package consisted of three Rapid SARS-CoV-2 Antigen self-tests (MP Biomedicals), two nose/throat swabs (FLOQSwabs, Copan), a tube with gelatin-lactalbumin-yeast (GLY) viral transport medium (Mediaproducts BV, Groningen, The Netherlands) for collecting a nose and throat sample, a Rigid Safety Bag (DaklaPack), and a Covermed medical return envelope (DaklaPack) to meet national posting criteria for medical diagnostic materials.

When participants were invited to send in a nose and throat sample, they could use the aforementioned materials along with a lab return slip. To ensure correct processing, an instruction sheet, and instruction video were available.

Participants were asked to send the nose and throat sample by mail on the same day the sample was collected, or when not possible due to weekend or isolation, keep the closed envelop in the fridge and post it on the earliest workday.

The packages were delivered in the weeks leading up to the start of the study on October 3, 2022, or within two weeks of registration after this date. Whenever a participant sent in a nose and throat swab, a new self-test package was dispatched.

Participants were free to request additional self-test packages at any time during the 2022/2023 season. Furthermore, simplified laboratory testing results (i.e. common cold, flu, COVID-19) were reported to participants in an online personal portal at a minimum of two weeks from the date of self-collection of the nose and throat sample. This two-week interval was applied to prevent any influence on participants’ healthcare-seeking behaviour and to ensure that the results are not perceived as a (timely) medical diagnosis.

### Inclusion to send in a (random) sample

We aimed to collect as many SARS-CoV-2 samples as possible to survey circulating variants of SARS-CoV-2 in the general population. Thus, all participants who reported a positive SARS-CoV-2 self-test within two days prior to filling in the questionnaire, irrespective of ARI symptoms, were invited to send in a nose and throat sample (Figure 1). Participants could be invited to send in a nose and throat swab multiple times during the study period.

**Figure 1.**
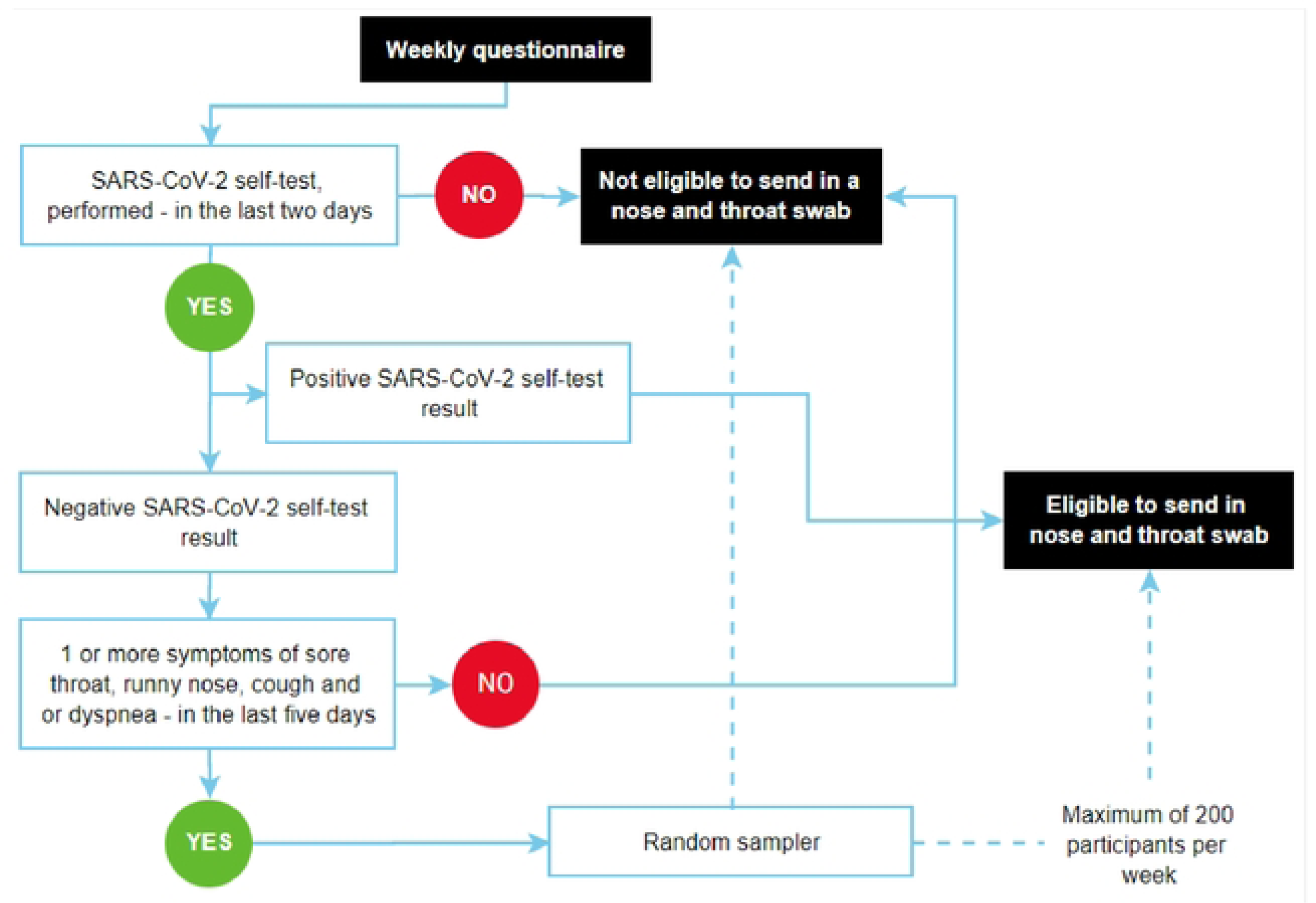
Sampling strategy flow chart.

In order to control laboratory workload and costs, a random sample of maximum 200 per week of those who reported a negative self-test while experiencing respiratory symptoms were selected to send in a nose and throat swab. Those who tested negative within two days prior to filling in the questionnaire and had a symptom onset (sore throat, runny nose, cough, or dyspnoea) within five days prior to completing the questionnaire, were potentially eligible for selection. (Figure 1).

To assure random sampling over the week we worked with a ticketing-system. First, before each week we sampled 200 exact time-slots (seconds after midnight Sunday-Monday), out of a dataset with 100.000 observed timings of submissions of weekly questionnaires (in seconds after midnight Sunday-Monday) in the 2021/2022 season. Secondly, when a time-slot is reached an additional ticket is made available and added to a stack. When a participant full-fills the inclusion criteria the system checks if a ticket is available in the stack. If available, the ticket is assigned to the participant. When the participant accepts the invitation, the ticket is removed from the stack. When the participant declines the invitation, the ticket goes back in the stack. If no ticket is available, the participant is not invited. At the end of the week the stack is reset to zero. Using this method we sample randomly over the time of the week, taking into account that most people participate in the evening, and slightly less on weekends, but more on Mondays, as these intricate patterns are included in the observed timings.

### Laboratory procedure

Nose and throat samples from participants with a positive SARS-CoV-2 self-test were analyzed for SARS-CoV-2 by RT-qPCR in the RIVM laboratory using the TaqMan® Fast Virus 1-Step Master Mix (Thermo Fisher) and Roche LC480 II thermal cycler with SARS-like beta coronavirus (Sarbeco) specific E-gene and SARS-CoV-2 specific RdRP primers and probe as described (7). SARS-CoV-2 positive samples with a cycle threshold value below 30 were sequenced to determine variants (8). Nose and throat samples from participants with a negative SARS-CoV-2 self-test were tested for 22 respiratory pathogens with multiplex-PCR of RespiFinder® 2Smart (PF2600-2S) (9) at the RIVM laboratory. In this multiplex-PCR rhinovirus/enterovirus are not sufficiently distinguishable, hence a follow-up PCR was performed on a sub-set to infer something about the contribution of rhinovirus and enterovirus.

### Analysis

We conducted a comprehensive analysis to examine various aspects of the study. First, we assessed participant demographics, questionnaire completion rates, the prevalence of ARI symptoms, and the number of received swabs. Stratification was performed based on gender, age group and region of residence. Participants who only filled in the intake questionnaire but did not fill in at least one weekly questionnaire were excluded from the analysis. Next, we analysed delay times in receiving nose and throat swabs at the RIVM laboratory. Subsequently, we investigated SARS-CoV-2 variant distributions and respiratory pathogen detections over time. We present results for the respiratory season 2022/2023, which is from October 3, 2022, to May 21, 2023 (week 40 2022-week 20 2023).

Nose and throat swab results from participants who refused to report their SARS-CoV-2 self-test result were excluded from the analysis. Additionally, swabs from participants who submitted their samples without being invited and did not have ARI symptoms or had a symptom onset longer than five days prior to completing the questionnaire were also excluded from the analysis.

The participatory surveillance software used in this study was developed in context of InfluenzaNet and is open source(10). Test results were reported back using proprietary software GLEAN (11). Data were analysed with R statistical software version 4.3.1 (12), with packages dplyr (13), ggborderline (14), ggplot2 (15), lubridate (16), readxl (17), tableone (18) and tibble

(19).

### Ethical considerations and privacy

The research protocol was shared with the Medical Ethics Review Committee Utrecht, and an official waiver for ethical approval (reference number: WAG/avd/20/008757; protocol 20-131) was obtained given the nature of data collection.

Participants had to agree to the privacy statement upon registration, which described the processing of personal data and research results, website security measures taken, and how to file a complaint. Additionally, they had to give consent to participate in the study. Participants were eligible to withdraw from the study at any time. Persons had to be 16 years or over to be able to participate.

## Results

### Participation

During the study period, a total of 17,030 individuals were included. This includes 8,302 participants recruited from existing users, 7,729 included via the random population sample, and 999 participants who joined during the season. The participants included based on the random sample reflect a response rate of 2.7% for the recruitment via the random population sample; for further details on these response rates, see Appendix 1. All participants combined completed 408,631 questionnaires in total (median: 29 per participant) over a span of 33 weeks. The median number of weeks that participants filled out at least one questionnaire was 26 weeks and 2,133 (12%) participants completed a weekly questionnaire each week over the entire study period. A total of 11,979 (70%) participants reported one or more times ARI symptoms with an onset of five days prior to completing the questionnaire. Consequently, 5,498 invitations were sent to participants meeting the inclusion criteria for submitting a nose and throat swab (*n*=4,582). Of these invitations, 4,062 (74%) were accepted and successfully followed up with a swab. A total of 782 (17 %) participants were asked more than once to submit a nose and throat swab, whereof 632 (81 %) did so. It should be noted that another 1,833 nose and throat samples were sent in without a formal invitation. Thereof, 1,509 (82.3%) came from participants having ARI symptoms and a symptom onset within five days prior to completing the questionnaire, but 324 (21.5%) of uninvited samples did not meet these criteria. These latter samples are excluded from further analysis.

Table 1 presents the characteristics of all participants, participants with ARI symptoms at least once during the study period and participants who sent in at least one nose and throat swab to the RIVM, alongside the characteristics of the overall population in the Netherlands. In the entire study population, males were underrepresented (42.7%) compared to females (57.1%). This gender disparity was more pronounced in the subgroup of participants with ARI symptoms as well as who sent in one or more nose and throat swabs. Also, there was a relative high participation level among participants aged 40 to 59 years (41.5%) and 60 to 79 years (27.8%) compared to the corresponding proportions in the general population of the Netherlands (31.9% and 14.8%, respectively). ARI symptoms were reported, and nose and throat samples were sent in most frequently by participants in the age group 40 to 59 years. Moreover, the spatial distribution of participants and subgroups appeared to align well with the geographical distribution observed in the Netherlands.

**Table 1.**
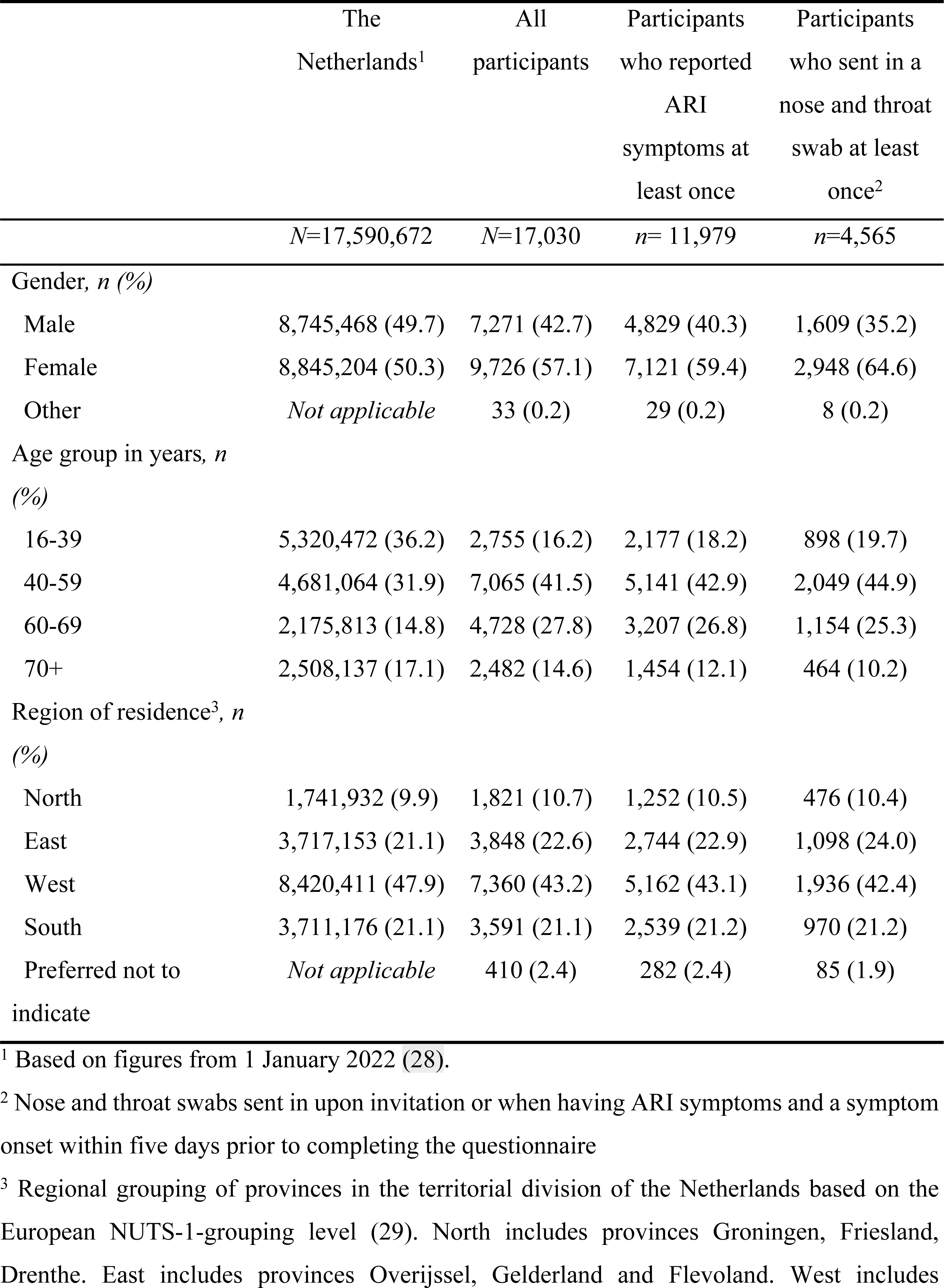
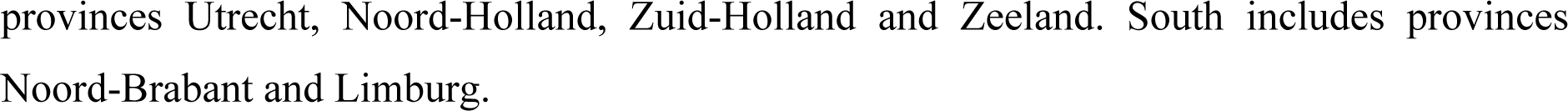
Characteristics of participants compared to the total population in the Netherlands.

|  | The Netherlands<sup>1</sup> | All participants | Participants who reported ARI symptoms at least once | Participants who sent in a nose and throat swab at least once<sup>2</sup> |
| --- | --- | --- | --- | --- |
|  | *N*=17,590,672 | *N*=17,030 | *n*= 11,979 | *n*=4,565 |
| Gender, *n* (%) |  |  |  |  |
| Male | 8,745,468 (49.7) | 7,271 (42.7) | 4,829 (40.3) | 1,609 (35.2) |
| Female | 8,845,204 (50.3) | 9,726 (57.1) | 7,121 (59.4) | 2,948 (64.6) |
| Other | *Not applicable* | 33 (0.2) | 29 (0.2) | 8 (0.2) |
| Age group in years, *n* (%) |  |  |  |  |
| 16-39 | 5,320,472 (36.2) | 2,755 (16.2) | 2,177 (18.2) | 898 (19.7) |
| 40-59 | 4,681,064 (31.9) | 7,065 (41.5) | 5,141 (42.9) | 2,049 (44.9) |
| 60-69 | 2,175,813 (14.8) | 4,728 (27.8) | 3,207 (26.8) | 1,154 (25.3) |
| 70+ | 2,508,137 (17.1) | 2,482 (14.6) | 1,454 (12.1) | 464 (10.2) |
| Region of residence<sup>3</sup>, *n* (%) |  |  |  |  |
| North | 1,741,932 (9.9) | 1,821 (10.7) | 1,252 (10.5) | 476 (10.4) |
| East | 3,717,153 (21.1) | 3,848 (22.6) | 2,744 (22.9) | 1,098 (24.0) |
| West | 8,420,411 (47.9) | 7,360 (43.2) | 5,162 (43.1) | 1,936 (42.4) |
| South | 3,711,176 (21.1) | 3,591 (21.1) | 2,539 (21.2) | 970 (21.2) |
| Preferred not to indicate | *Not applicable* | 410 (2.4) | 282 (2.4) | 85 (1.9) |

<sup>1</sup> Based on figures from 1 January 2022 (28).
<sup>2</sup> Nose and throat swabs sent in upon invitation or when having ARI symptoms and a symptom onset within five days prior to completing the questionnaire
<sup>3</sup> Regional grouping of provinces in the territorial division of the Netherlands based on the European NUTS-1-grouping level (29). North includes provinces Groningen, Friesland, Drenthe. East includes provinces Overijssel, Gelderland and Flevoland. West includes
provinces Utrecht, Noord-Holland, Zuid-Holland and Zeeland. South includes provinces Noord-Brabant and Limburg.

### Collection of nose and throat samples

The 4,565 participants who contributed one or more nose and throat swabs, sent in a total of 5,571 swabs, comprising 1,475 (median: 37 per week) samples from participants with a positive SARS-CoV-2 self-test result and 4,096 (median: 136 per week) samples from participants with a negative SARS-CoV-2 self-test result.

Table 2 presents the median and interquartile range (IQR) times for participants’ SARS-CoV-2 results in relation to the first symptom onset, performing a SARS-CoV-2 self-test and self-swabbing a nose and throat sample. The median time between the first symptom onset and performing a SARS-CoV-2 self-test was 1.0 day (IQR= [1.0, 2.0]) for both the positive and negative self-test groups. Similarly, the median time between performing a SARS-CoV-2 self-test and self-swabbing the nose and throat sample was 1.0 day (IQR= [0.0, 2.0]) for both groups. Furthermore, the median time between the first symptom onset and self-swabbing the nose and throat sample for both groups was 2.0 days (IQR= [1.0-3.0]). Participants with a positive SARS-CoV-2 self-test result had a median time of 4.0 days (IQR=3.0-6.0) between the first symptom onset and for the RIVM laboratory to register the nose and throat sample, while those with a negative self-test result this was 5.0 days (IQR=3.0-6.0). The availability of the self-test package at home, before start of symptoms, allowed for these short intervals.

**Table 2.** Delay times in days between first symptom onset, performing a SARS-CoV-2 self-test and self-swabbing a nose and throat sample. For samples coming from positive SARS-CoV-2 self-tests and for samples coming from negative SARS-CoV-2 self-tests.

|  | Positive<br>SARS-CoV-2<br>self-test result<br>*n*=1,159 | Negative<br>SARS-CoV-2<br>self-test result<br>*n*=3,736 |
| --- | --- | --- |
| Time between first symptom onset and performing a SARS-CoV-2 self-test in days, median (IQR) | 1.0 [1.00, 2.00] | 1.0 [1.00, 2.00] |
| Time between performing a SARS-CoV-2 self-test and self-swabbing nose and throat sample in days, median (IQR) | 1.0 [0.00, 2.00] | 1.0 [0.00, 2.00] |
| Time between first symptom onset and self-swabbing nose and throat sample in days, median (IQR) | 2.00 [1.00, 3.00] | 2.00 [1.00, 3.00] |
| Time between first symptom onset and nose and throat sample registration date at the RIVM laboratory in days, median (IQR) | 4.00 [3.00, 6.00] | 5.00 [3.00, 6.00] |
| Median (interquartile range (IQR)). |  |  |

### Outcomes of virological analysis

The positive SARS-CoV-2 self-test result could be confirmed in 1417 (96.1%) of the nose and throat samples. Negative SARS-CoV-2 self-test results were found to be SARS-COV-2 positive in 295 (7.2%) of the nose and throat samples using PCR, likely related to differences in sensitivity and/or sampling.

Figure 2 shows the distribution of SARS-CoV-2 genetic clades within positive samples at the RIVM laboratory and the national SARS-CoV-2 variant surveillance (20) on a weekly basis. Within the samples from the RIVM laboratory the clades 22B (Omicron, BA.5), 22E (Omicron, BQ.1) and 23A (Omicron, XBB.1.5) and related sub-variants predominated throughout the study period with an average of 20.2%, 25.4% and 25.7% respectively. Among these, 22B had the highest peak with 81.4% in year-week 2022-40, accounting for almost all samples in this week. These patterns are consistent with findings in the national SARS-CoV-2 variant surveillance (20).

**Figure 2.**
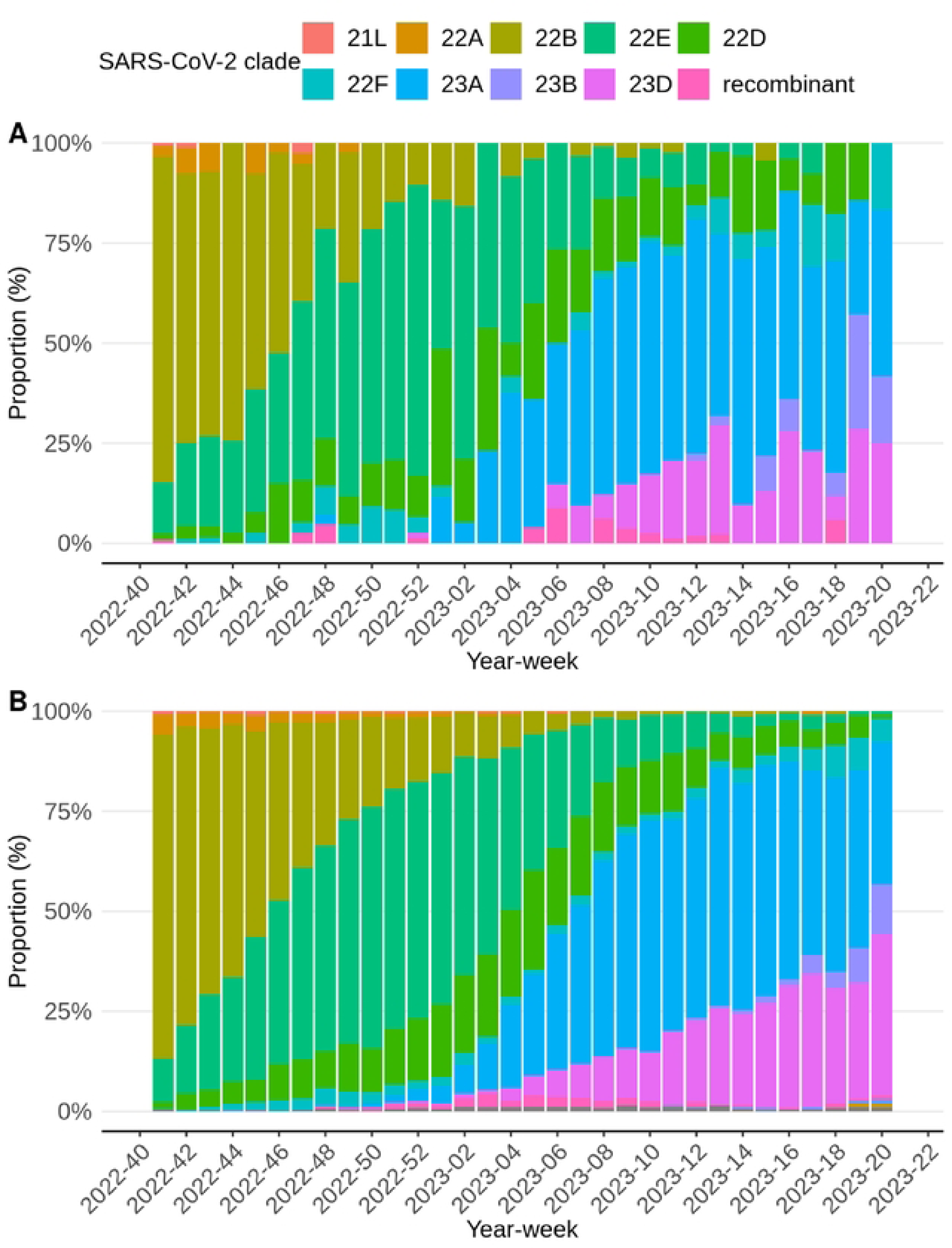
Proportional SARS-CoV-2 clade distributions per week of Infectieradar (A) and national pathogen surveillance in the Netherlands (B).

Samples from participants with a negative SARS-CoV-2 self-test (n = 4,096), were tested for 22 respiratory pathogens. Of those samples 1,955 (47.7%) tested positive for one or more of the tested pathogens. Figure 3 shows the distribution of pathogens tested positive for among those samples, per week, starting from week 40 in 2022. Some samples contained multiple pathogens and are therefore included multiple times in this figure. The number of weekly samples varied from 45 to 191 (median 136). The percentage of positive samples dropped temporarily the last weeks of 2022 and the first weeks of 2023 and consistently from week 11 2023 onwards. During the whole study period rhinovirus/enterovirus accounted for the majority of the respiratory pathogens identified, with an average of 44.5%. Follow up PCR based typing on a representative sample subset revealed that approximately 98% of these were Rhinovirus infections with less than 2% Enterovirus detections. 7% tested positive for SARS-CoV-2 in the PCR despite a negative antigen-based self-test. Other important respiratory pathogens like seasonal coronaviruses, Influenza virus A and B, RSV-A and RSV-B and hMPV were also detected in our study population.

**Figure 3.**
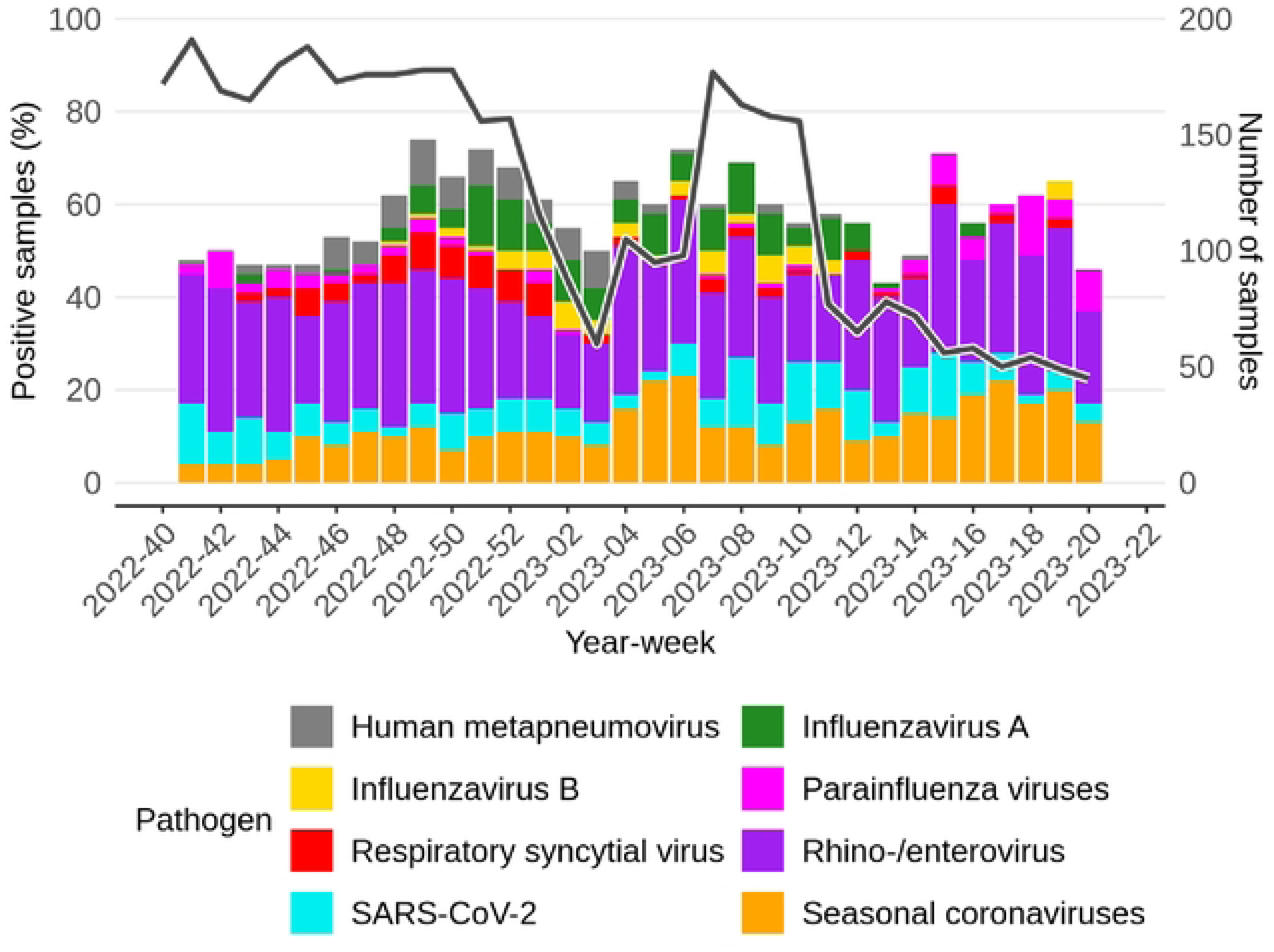
Samples taken from participants with symptoms associated with an acute respiratory infection and with a negative SARS-CoV-2 self-test result per week. The colors indicate the pathogens the samples tested positive for. Some samples were tested positive for more than one pathogen and are counted for all pathogens. The black solid line indicates the number of samples, that can be seen on the y-axis on the right.

To provide an estimate of the pathogen-specific incidence of ARI throughout the season 2022/2023, the percentage of participants experiencing ARI symptoms was multiplied with the percentage of positive samples tested at the RIVM laboratory per pathogen per week as shown in Figure 4. Notably, rhino-/enterovirus incidence shows the highest values in nearly all weeks, starting with a peak at year-week 2022-40. Moreover, while seasonal coronaviruses exhibited significantly lower values in comparison to rhino-/enterovirus, they also displayed a similar pattern, with peaks occurring during year-weeks 2022-47 and 2023-06. Note, due to our sampling strategy SARS-CoV-2 in this graph is based on those who had a negative self-test. Therefore we also show the overall reports of positive SARS-CoV-2 self-tests in Figure 5.

**Figure 4.**
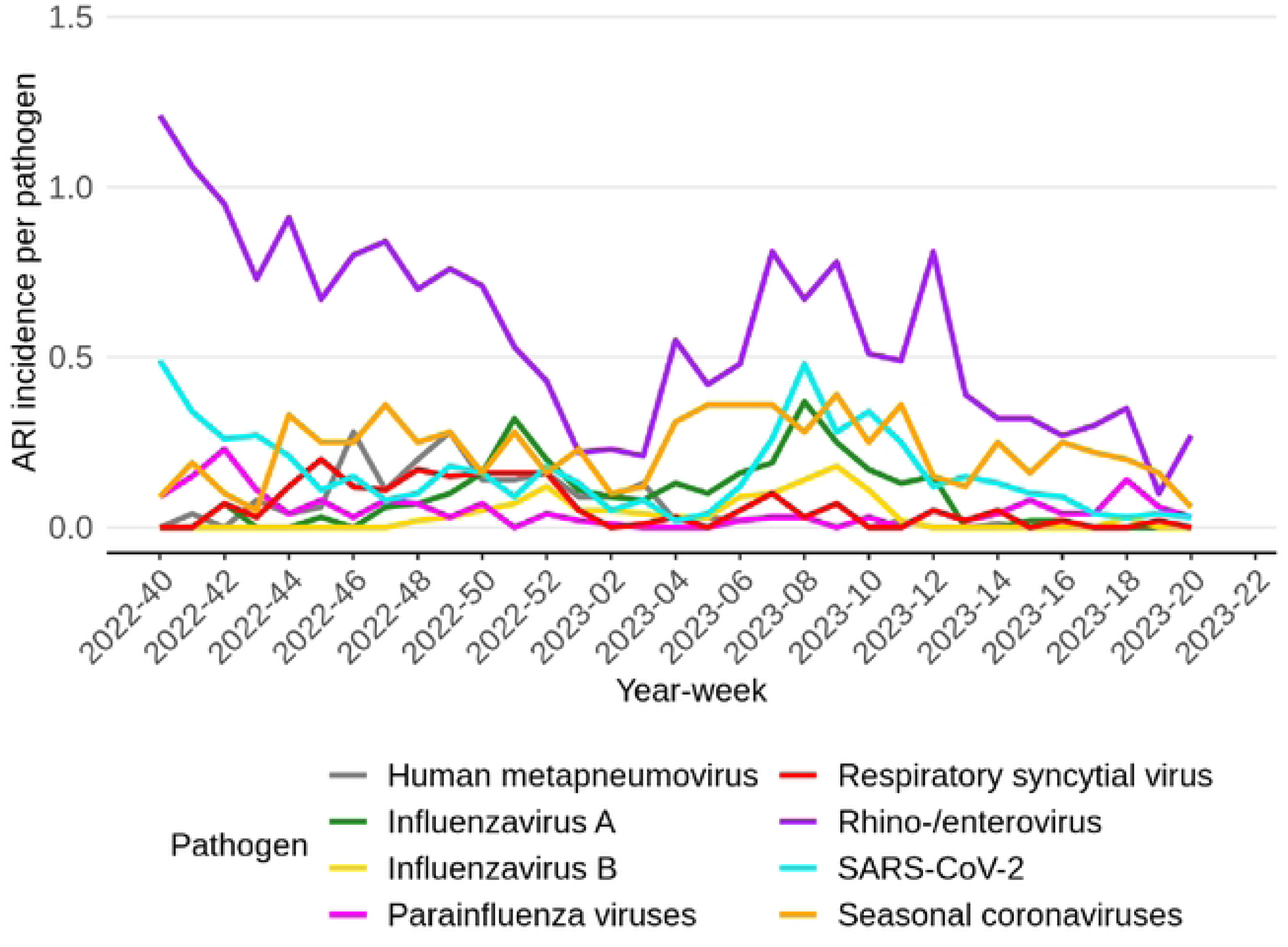
Samples taken from participants with symptoms associated with an acute respiratory infection and with a negative SARS-CoV-2 self-test result per week. The line indicates the ARI incidence multiplied by percentage of samples that were tested positive for the respective pathogen, which are indicated by color. Note, due to our sampling strategy SARS-CoV-2 in this graph is based on those who had a negative self-test

**Figure 5.**
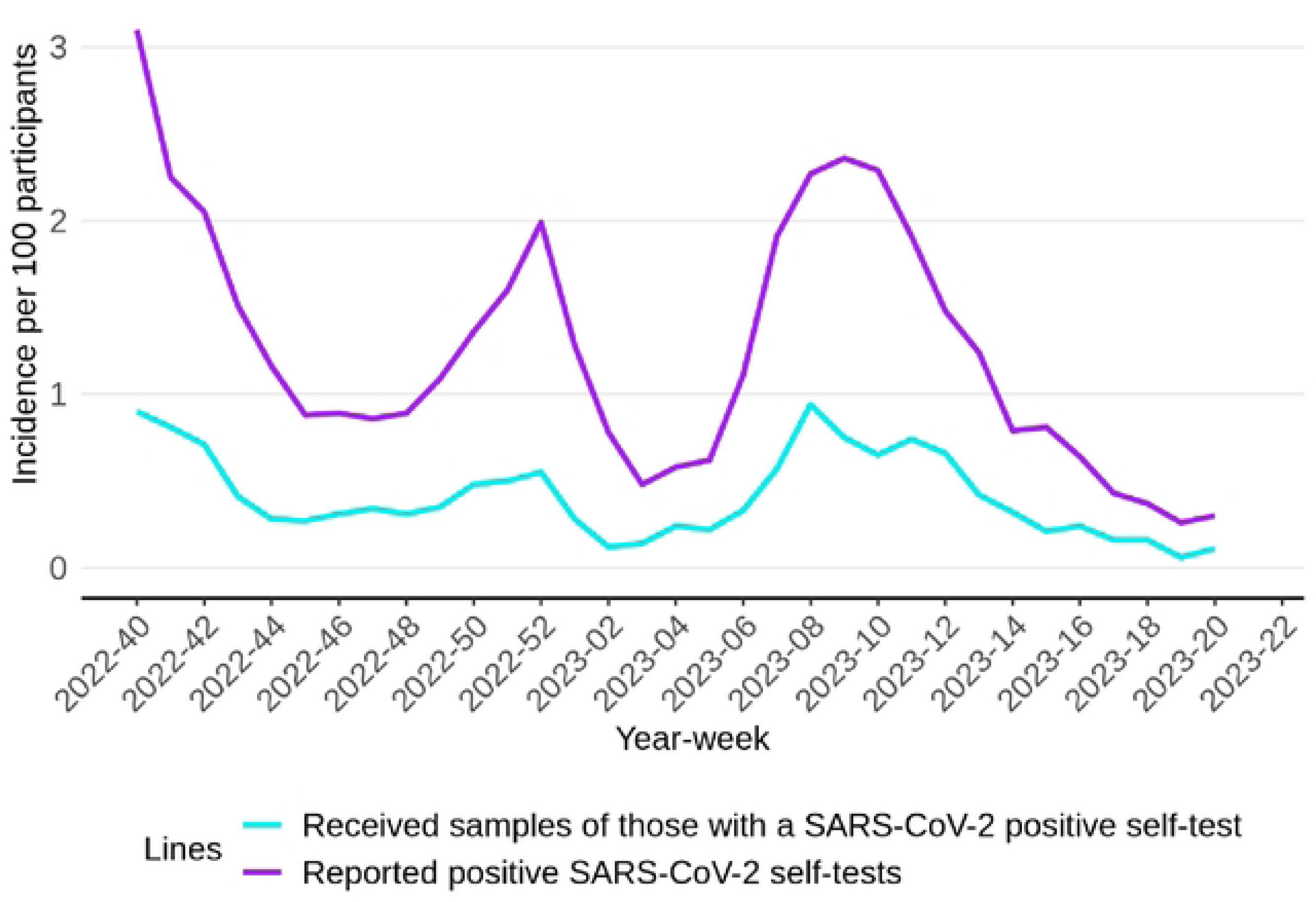
Reported positive SARS-CoV-2 self-tests per week, as well as the received nose­ and throats samples of those with a positive self-test.

## Discussion

Here we show that it is meaningful and feasible to run a large online participatory syndromic surveillance system which includes centralized virological testing using self-swabs. The data was collected timely and the system was sustainable over the full season. Therefore, we see this as a powerful new tool for the near real time surveillance of infectious diseases, inside and outside pandemic times. It combines the molecular insight of virological surveillance with the flexibility and scale of participatory syndromic surveillance. We think that our example could serve others who wish to deploy such a system in their own country.

Others have experimented before with the combination of participatory surveillance and the collection of samples. Before the SARS-CoV-2 pandemic, in the 2011/2012 season such a study was run in Sweden among 2,237 participants (21), in the 2013/2014 kits were provided among 294 participants of a program in the US (22), and among the participants of FluSurvey in the UK (23) a pilot with self-swabbing including testing for influenza virus was performed. Also in the UK a multi-year household cohort Flu Watch (24) included the collection of nasal samples by post, tested for influenza virus. During the SARS-CoV-2 pandemic large cohorts were set up, such as the COVID-19 Infection survey in the UK which did include the collection of samples and testing for SARS-CoV-2 (25). Hence, although our approach is similar to what has done before, to our knowledge our system is the first which combines national participatory syndromic surveillance with broader virological testing, and integrates such system within overall national infectious disease surveillance.

The added value of this combined system, at scale, is manyfold. There is the ability to contribute to the early detection of outbreaks, monitor circulating pathogens and their variants, disease severity, evaluate interventions, identification of risk-groups, and contribute to more in-depth research to enable to subsequently inform public health policy and resource allocation. The possible contribution to research should not be undervalued as our understanding of basic processes in the evolution of pathogens is still limited. For example there are important outstanding questions on the interplay between enhanced transmission and virulence, in relation to population immunity (including vaccination), specific host-factors and behavioral differences (26, 27). The long-term and weekly follow-up in our cohort, including sampling, allows for an elaborate study design including nested studies. Furthermore, given that it is participatory surveillance, including volunteers, and timely and transparent reporting, our approach may contribute to the public trust in surveillance. And, when adapted on a larger scale, for example across Europe, we are confident such a network could contribute to international aims of global health security.

We feel that our approach delivers value for money. Our system’s overall costs are likely lower compared to alternative respiratory surveillance methods. Furthermore, as we know our participants, it is relatively cheap to add additional studies, because finding and recruiting participants is a resource intensive process, and the additional effort for the participant is also limited.

An important current limitation is our participant composition. Although our cohort includes participants from all parts of the country, our participants are not a reflection of the Dutch population. Younger age groups are missing. Currently, participants hardly include children attending nursery, primary, secondary school and only to a minor degree those attending higher education. Furthermore, although we don’t actively collect information on this, minority communities probably are underrepresented in our cohort and our cohort could include clusters of closely related participants. Such clusters could enhance biases in our sampling. However, our collected SARS-CoV-2 genetic data shows that the variant distribution over time is very comparable with the distribution in the national variant surveillance, indicated little to no disturbing effects or biases of allowing inclusion of multiple household members. Therefore, our cohort offers meaningful insights into what is circulating in the wider community. Varieties in the variant distribution follows the national pattern in our cohort just as in the national surveillance, albeit sometimes a little later or earlier, and in a slightly different frequency. This is an important validation of the contribution of our cohort. In the current study existing participants can add their children to our survey, but only a small minority does so. It will be therefore challenging to improve this towards the short-term future, unless there is an outbreak situation when there is an urgent need to do so, allowing more active recruitment. Therefore, we are exploring school-based methodologies to improve syndromic surveillance among those too young to participate. In the future, we aim to improve the participation of minority groups by offering our questionnaires and communication in multiple languages.

We experienced also operational problems. With a response rate of 2.6% our recruitment effort was inefficient. This response is based on one recruitment letter, without reminders, and without supporting communication effort. Adding a reminder or improved communication would likely have improved the response. A certain fatigue related to COVID-19 might have been present in the population during recruitment in September 2022. In case of a new emerging infection, we aim to recruit larger numbers in the early phases of the outbreak. In contrast, some of those recruited did more than requested and sent in samples when not explicitly asked for. Although this led to more samples in our study, it made the samples were selected less random and led to more costs. Furthermore, around the end of 2022 and the start of 2023 we saw a drop in the number of samples. This drop was linked to a software-bug in the algorithm which invites participants to send in a sample. The bug prevented participants from being invited to submit samples a second time even though they reported new symptoms. In addition, around Christmas 2022 we did experience a delay in samples coming in due to seasonal pressures on the postal service. And we had some login problems linked to the expiration of accounts in the software we use to provide lab-results to the participants. Although some of these problems took effort to solve, all in all they were minor and did not impact on the integrity or utility of our results.

A strength of our study was the high compliancy rate of participants to send in a sample when we asked to do so, and the short delay between onset and collection of the sample due to the fact that sampling kits are present at the participants before start of symptoms. We believe that our approach to provide every participant both antigen tests and nose- and throat swabs at the start of the study instead of sending test-kits when participants report symptoms contributed to this.

We developed our surveillance system as part of a national public health strategy because participatory syndromic surveillance focusses on one layer of the surveillance pyramid (symptomatic disease without (or before) need of health care) and therefore integration with data from other surveillance systems is essential to obtain a complete picture of the disease burden and the distribution of pathogen associated with complaints.

We consider our observations a proof of principle of such centralized system relevant for the surveillance toolbox and pandemic planning. As the additional testing doesn’t need to be up and running every season and could be deployed and scaled up quickly when deemed necessary – actually in the season 2023/2024 our inclusion changed to focus more on acute respiratory infections in general instead of a focus on SARS-CoV-2. Furthermore, collected samples don’t need to be tested each week but can be tested batch-wise which can be less frequent, or more stringent selection-criteria (e.g. severity of symptoms) could be defined before inviting someone to send in a swab. In case of a new emergent disease, samples can be collected, stored and tested later when pathogen specific tests are not available yet, while data can be collected about symptoms, duration of illness and other endpoints.

Our method includes many parts of a citizen science philosophy. We provided test results to the participants and made insights in the data available in real time via weekly updates of our data dashboard and publication on the national surveillance website. Towards the future we could improve on including participants in the definition of the research questions, and include them more explicitly in data-analytics, modelling and interpretation. We do believe that the citizen science model can improve trust in our findings and conclusion and also improve long term recruitment. Therefore, we aim to expand the role of our participants.

In conclusion, this large-scale, centralized participatory surveillance system provides a comprehensive approach for performing syndromic and virological surveillance in the general population, including respiratory pathogen detection by self-test or multiplex-PCR. The continuous collection of samples among those who don’t seek care the system is part of an answer on how to study the transmission, competition, virulence and evolution of circulating pathogens in interpandemic periods.

## Data Availability

Not all data cannot be shared publicly because of legal constrains. Data underlying the results are also available on www.infectieradar.nl/results and data requests can be send to.

## Abbreviations

ARI: Acute Respiratory Infection
BRP: Basis Registratie Persoonsgegevens
COVID-19: coronavirus disease
GP: general practitioner
ILI: influenza like illness
IQR: interquartile range
RIVM: Rijksinstituut voor Volksgezondheid en Milieu SARS-CoV-2 Severe Acute Respiratory Syndrome Coronavirus 2

